# Electrocardiographic signs of cardiac ischemia at rest and during exercise in patients with COPD travelling to 3100 m. Data from a randomized trial of acetazolamide

**DOI:** 10.1101/2024.11.12.24317157

**Authors:** Marla Christen, Aline Buergin, Maamed Mademilov, Laura Mayer, Simon R. Schneider, Mona Lichtblau, Talant M. Sooronbaev, Silvia Ulrich, Konrad E. Bloch, Michael Furian

## Abstract

**Introduction:** In patients with chronic obstructive pulmonary disease (COPD), oxygen delivery to the heart may be impaired during altitude travel. We assessed ECG-derived signs of cardiac ischemia, and effects of preventive acetazolamide therapy in COPD patients travelling to high altitude.

**Methods:** Patients with COPD, GOLD grade 2-3, mean±SD, FEV_1_ 66±11%predicted, aged 57±8years, living <1000m, were included in this analysis of secondary outcomes from a randomized, placebo-controlled, double-blind trial (www.clinicaltrials.gov, NCT03156231). Exercise-electrocardiograms were recorded at the National Center of Internal Medicine and Cardiology, Bishkek (760m) and on the day of arrival at Tuja Ashu high-altitude clinic (3100m), Kyrgyzstan. Acetazolamide (375mg/day) or placebo was administered 24h before ascent and during stay at 3100m. The incidence of post-exercise ST-elevations (STE) ≥0.3mm in aVR (J+80ms) was the main outcome.

**Results:** At 760m, 3 of 49 (6%) patients randomized to placebo and 3 of 50 (6%) randomized to acetazolamide showed post-exercise STE. At 3100m under placebo, 2(4%) new STE developed and 1(2%) disappeared compared to 760m (P=0.564, McNemar Test). At 3100m under acetazolamide, 1(2%) new STE developed and 2(4%) disappeared compared to 760m (P=0.564). No treatment effect was detected (P=0.242, Fisher Exact Test). Mean difference (95%CI) in STE between post-exercise and rest at 3100m was 0.22mm(0.06 to 0.39) and 0.09mm(−0.06 to 0.24) under placebo and acetazolamide therapy (treatment effect, −0.13mm(−0.35 to 0.08, P=0.230)).

**Conclusions:** In lowlanders with moderate to severe COPD ascending to 3100m, no ECG-derived signs of cardiac ischemia emerged neither at rest nor post-exercise and this was not modified by preventive acetazolamide therapy.

## Introduction

Chronic obstructive pulmonary disease (COPD) has a global prevalence of 8-15%, is ranked 9^th^ among causes of years of life lost, and is expected to rise to the 4^th^ rank by 2040.(1) Patients with COPD suffer from ventilatory and pulmonary gas exchange impairments associated with chronic hypoxemia, which may lead to pulmonary hypertension or ischemic heart disease. The main symptoms of COPD are chronic cough, dyspnea and exercise impairment near sea level. Due to its high prevalence in the general population, COPD is also expected to be common among mountain tourists.

At high altitude, the reduced barometric pressure and lower inspiratory partial pressure of oxygen exacerbate hypoxemia in patients with COPD and they may promote right heart impairments and cardiac repolarization disturbances.(2–7) At low altitude, recent research showed that post-exercise elevation in the ST-segment in aVR of ≥0.3 mm (STE) is a robust predictor of myocardial ischemia independent of ST depressions in V5 and clinical factors.(8) However, reports on ECG changes in COPD at high altitude are scant. They include subclinical prolongations in the QT-interval(9) and reductions in the ST-segment in V5 at rest and during exercise.(10) Furthermore, effects of preventive treatment with acetazolamide, a drug used in prevention of acute mountain sickness in healthy individuals, have not been studied.(9, 10)

Therefore, the purpose of this study was to test the hypotheses that patients with COPD will develop signs for cardiac ischemia in the aVR when travelling to high altitude and that this may be prevented by acetazolamide.

## Methods

### Study design and setting

This trial evaluating ECG signs of cardiac ischemia was nested within a randomized, placebo-controlled, double-blind, parallel design trial evaluating the effect of preventive acetazolamide therapy on altitude-related adverse health effects (ARAHE) in patients with COPD during a stay at high altitude.(11) Cardiopulmonary exercise data have been published previously,(12) however, data related to ECG and cardiac ischemic have not been previously published. The study was conducted between May 01, 2017 and July 31, 2018. Baseline evaluations were performed at the National Center of Internal Medicine and Cardiology (760 m), Bishkek, Kyrgyzstan. Subsequently, patients were transferred with a minibus to the high-altitude clinic Tuja Ashu, Kyrgyzstan, located at 3100 m. Acetazolamide (375 mg/day, 125 mg in the morning and 250 mg in the evening) or equally looking placebo capsules were orally administrated one day before departure and during a 2-day sojourn at 3100 m according to a 1:1 randomization. The trial was approved by the Ethics Committee of the NCCIM (08-2016) and registered at www.clinicaltrials.gov (NCT03156231). Participants gave written, informed consent.

### Participants

Men and women, aged 18-75 years, living below 800 m, diagnosed with COPD according to the Global Initiative for Chronic Obstructive Pulmonary Disease (GOLD) guidelines,(13) post-bronchodilator forced expiratory volume in the first second (FEV_1_)/forced vital capacity (FVC)<0.7, FEV_1_ 40-80% predicted, pulse oximetry (SpO_2_) ≥92%, PaCO_2_<6.0 kPa) at 760 m were included (n=99). Exclusion criteria were COPD exacerbation, severe or unstable comorbidities and allergy to sulphonamides.

### Assessments

Examinations included weight, height, blood pressure, heart frequency, cardiac and pulmonary auscultation of the patients at 760 m and 3100 m.

### Exercise testing

At 760 m and within 4 hours of arrival at 3100 m, patients underwent cardiopulmonary exercise tests using a progressive maximal ramp protocol(14) starting at 20 W and increasing by 10 W/min to exhaustion. A 4-lead ECG was continuously recorded (AMEDTEC ECGpro) and the rolling 30 sec mean of the ST-segment amplitude in lead aVR was computed at 80 ms and 60 ms from the J point. Values from rest, peak exercise and shortly after exercise termination (post-exercise) were extracted.

Based on previous a recent study, providing a clinically relevant cut-off of ST-segment elevation in aVR of ≥0.3 mm,(8) the main outcome of interest in this study were the altitude and acetazolamide-related STE occurrences in aVR during post-exercise.

Further outcomes included exercise work rate, heart rate, blood pressure and pulse oximetry, as well as arterial blood gases obtained at peak exercise.

### Randomization

Participants were randomly allocated to either acetazolamide or placebo capsules by the software MinimPy.(15) Drug assignment followed a 1:1 allocation ratio as per computer generated schedule minimizing for age (18-50 and 51-75 years), sex and severity of airflow obstruction (FEV_1_ 40-59 and 60-80% predicted). An independent pharmacist prepared active or identically looking placebo capsules labelled with concealed codes. Patients and researchers remained blinded until data analysis was completed.

### Statistical analysis

The statistical analysis was perfomed with R (Version: 4.4.0.). Based on the Shapiro-Wilk test, the data showed a normal distribution and a per-protocol analysis was performed on all available data. The results are presented as mean ± SD and mean differences (95% confidence intervals). Altitude-induced changes in STE prevalence ≥0.3 mm in aVR were compared by the McNemar test accounting for paired comparisons. The treatment effect of acetazolamide was evaluated by Fisher’s exact test. A comparison was considered statistically significant with a p-value < 0.05.

## Results

Of 386 patients assessed for eligibility, 176 were randomized to either placebo (N = 90) or acetazolamide (N = 86). Of these, 77 randomized patients underwent no ECG recordings for various reasons (Figure 1). Therefore, the per-protocol analysis included data of 49 and 50 patients, randomized to placebo or acetazolamide.

**Figure 1:**
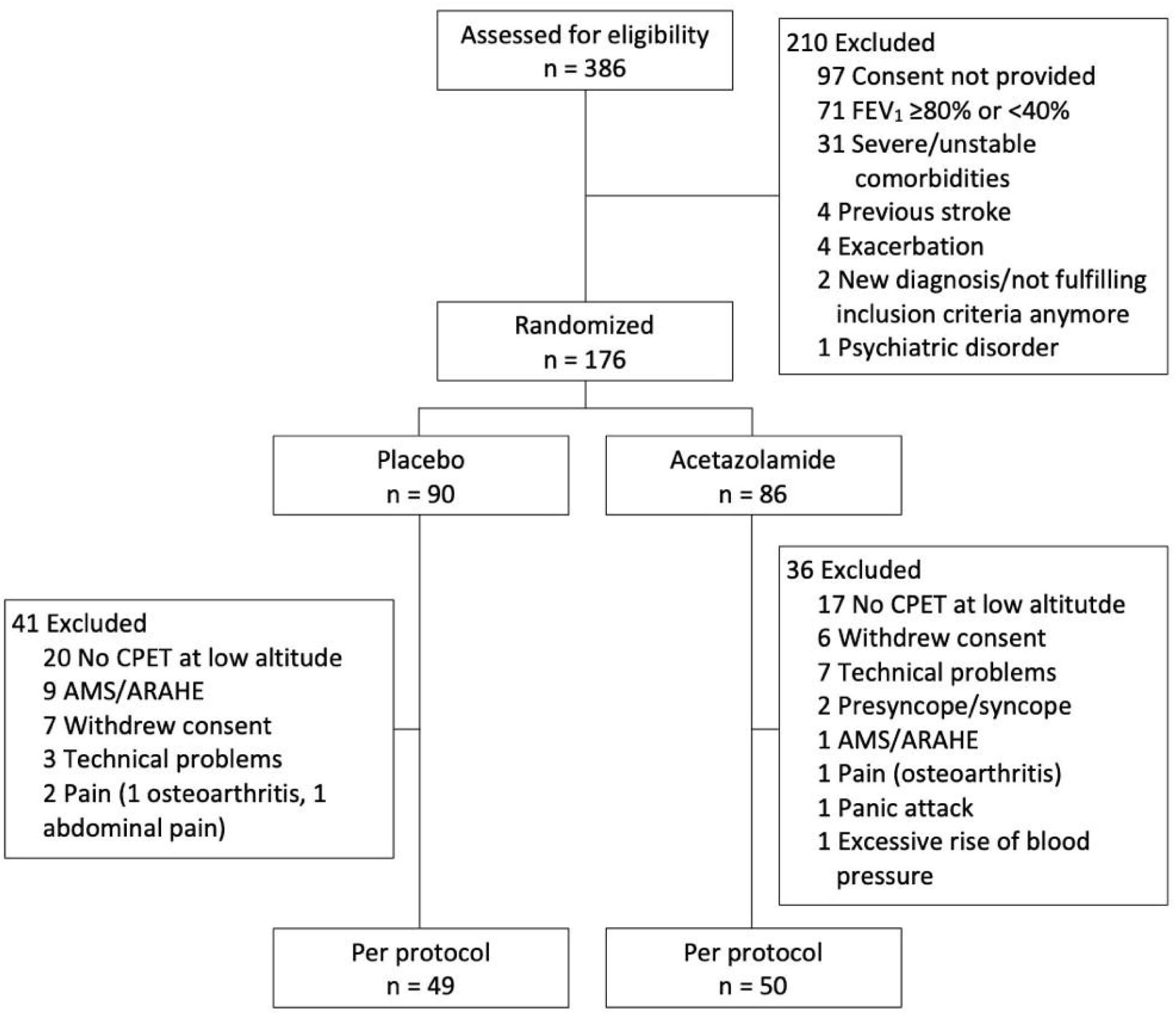
Study participant flow. AMS, acute mountain sickness; ARAHE, altitude-related adverse health effects; CPET, cardiopulmonary exercise testing; FEV_1_, forced expiratory volume in 1 second.

Patient characteristics are presented in Table 1. The age was, mean ± SD, 57.0 ± 9.7 years for the placebo and 57.5 ± 6.6 years for the acetazolamide group. The majority of the patients (89%) were diagnosed with COPD GOLD grade 2.

**Table 1:** Patient characteristics.

| Characteristic | Placebo group | Acetazolamide group |
| --- | --- | --- |
| No (women) | 49 (13) | 50 (18) |
| Age, year | 57.0 ± 9.7 | 57.5 ± 6.6 |
| Height, m | 1.64 ± 0.08 | 1.65 ± 0.09 |
| Weight, kg | 72.7 ± 15.2 | 74.4 ± 12.6 |
| BMI, kg/m<sup>2</sup> | 27.1 ± 5.0 | 27.2 ± 3.8 |
| FEV<sub>1</sub>, % predicted | 66 ± 12 | 65 ± 10 |
| FEV<sub>1</sub>/FVC | 0.60 ± 0.09 | 0.62 ± 0.08 |
| GOLD grade (2/3) | (42/7) | (46/4) |
| mMRC, n (%) |  |  |
| 0 | 27 (55.1) | 29 (58.0) |
| 1 | 14 (28.6) | 12 (24.0) |
| 2 | 4 (8.2) | 4 (8.0) |
| 3 | 3 (6.1) | 3 (6.0) |
| 4 | 1 (2.0) | 2 (4.0) |
| **Regular medication, n(%)** |  |  |
| Oral steroid | 1 (2.0) | 0 (0.0) |
| Inhaled beta adrenergics | 8 (16.3) | 11 (22.0) |
| Inhaled anticholinergics | 10 (20.4) | 9 (18.0) |
| Inhaled corticosteroids | 5 (10.2) | 8 (16.0) |
| Antihypertensives | 4 (8.2) | 7 (14.0) |
| Aspirin | 4 (8.2) | 7 (14.0) |

Values presented as mean ± SD or No. (%). BMI, body mass index; FEV<sub>1</sub>, forced expiratory volume in 1 second; FVC, forced vital capacity; GOLD, global initiative for chronic obstructive lung disease; mMRC, modified Medical Research Council dyspnea score.

### Electrocardiogram

The ST-segment in lead aVR at 80 ms from J point (J + 80 ms) revealed a slight but statistically significant change in the placebo group when comparing STE at 3100 m to 760 m at post-exercise, mean difference (95%CI) of 0.22 mm (0.06 to 0.39, p<0.05). However, in all other measurements at J + 80 ms or J + 60 ms, there were no significant changes in the ST-segment in aVR neither due to altitude nor due to treatment with acetazolamide (Table 2).

**Table 2:** ST amplitudes in lead aVR and clinical evaluation.

| Variable | Placebo | | $\Delta$ Mean difference<br>(3100 m-760 m)<br>(95% CI) | Acetazolamide | | $\Delta$ Mean difference<br>(3100 m-760 m)<br>(95% CI) | Treatment effect<br>(95% CI) | P-value |
| --- | --- | --- | --- | --- | --- | --- | --- | --- |
|  | 760 m | 3100 m |  | 760 m | 3100 m |  |  |  |
| HR peak, bpm | 135 ± 3 | 134 ± 3 | -1 (-6 to 5) | 135 ± 3 | 129 ± 3 | -6 (-11 to 0) | -5 (-12 to 3) | 0.227 |
| Maximal load, watts | 107 ± 4 | 97 ± 4 | -11 (-14 to -7) | 104 ± 4 | 90 ± 4 | -14 (-17 to -11) | -3 (-8 to 1) | 0.140 |
| SpO<sub>2</sub> peak, % | 93.9 ± 0.5 | 84.2 ± 0.5 | -9.7 (-10.5 to -9.0) | 93.7 ± 0.5 | 85.9 ± 0.5 | -7.7 (-8.5 to -7.0) | 2.0 (1.0 to 3.1) | 0.002 |
| PaO<sub>2</sub> peak, kPa | 10.1 ± 0.1 | 7.6 ± 0.1 | -2.5 (-2.7 to -2.3) | 9.8 ± 0.1 | 8.1 ± 0.1 | -1.7 (-1.9 to -1.6) | 0.8 (0.5 to 1.1) | <0.001 |
| PaCO<sub>2</sub> peak, kPa | 5.2 ± 0.1 | 4.8 ± 0.1 | -0.4 (-0.5 to -0.3) | 5.1 ± 0.1 | 4.4 ± 0.1 | -0.7 (-0.8 to -0.6) | -0.3 (-0.5 to -0.2) | <0.001 |
| ST in aVR with j + 60ms, mm |  |  |  |  |  |  |  |  |
| at rest | -0.35 ± 0.06 | -0.30 ± 0.06 | 0.05 (-0.06 to 0.17) | -0.29 ± 0.06 | -0.30 ± 0.06 | 0.00 (-0.12 to 0.12) | -0.05 (-0.22 to 0.11) | 0.526 |
| at peak | -0.18 ± 0.06 | -0.10 ± 0.06 | 0.08 (-0.04 to 0.20) | -0.20 ± 0.06 | -0.21 ± 0.06 | -0.01 (-0.13 to 0.11) | -0.09 (-0.26 to 0.07) | 0.273 |
| post peak exercise | -0.44 ± 0.06 | -0.32 ± 0.06 | 0.12 (0.00 to 0.25) | -0.33 ± 0.06 | -0.31 ± 0.06 | 0.02 (-0.10 to 0.14) | -0.10 (-0.27 to 0.08) | 0.269 |
| Change rest to peak exercise | 0.17 ± 0.06 | 0.20 ± 0.06 | 0.03 (-0.13 to 0.20) | 0.09 ± 0.06 | 0.08 ± 0.06 | -0.01 (-0.17 to 0.15) | -0.04 (-0.27 to 0.19) | 0.711 |
| Change rest to post peak exercise | -0.09 ± 0.06 | -0.02 ± 0.06 | 0.07 (-0.10 to 0.24) | -0.04 ± 0.06 | -0.01 ± 0.06 | 0.03 (-0.14 to 0.19) | -0.05 (-0.28 to 0.19) | 0.919 |
| ST in aVR with j + 80ms, mm |  |  |  |  |  |  |  |  |
| at rest | -0.49 ± 0.07 | -0.40 ± 0.07 | 0.09 (-0.05 to 0.23) | -0.41 ± 0.07 | -0.38 ± 0.07 | 0.03 (-0.12 to 0.17) | -0.06 (-0.27 to 0.14) | 0.543 |
| at peak | -0.48 ± 0.07 | -0.34 ± 0.07 | 0.14 (-0.01 to 0.29) | -0.54 ± 0.07 | -0.41 ± 0.07 | 0.12 (-0.02 to 0.27) | -0.02 (-0.22 to 0.19) | 0.882 |
| post peak exercise | -0.79 ± 0.08 | -0.57 ± 0.08 | 0.22 (0.06 to 0.39) | -0.69 ± 0.07 | -0.60 ± 0.07 | 0.09 (-0.06 to 0.24) | -0.13 (-0.35 to 0.08) | 0.230 |
| Change rest to peak exercise | 0.01 (-0.14 to 0.16) | 0.06 (-0.09 to 0.20) | 0.05 (-0.16 to 0.25) | -0.13 (-0.27 to 0.01) | -0.03 (-0.18 to 0.11) | 0.10 (-0.11 to 0.30) | 0.05 (-0.24 to 0.34) | 0.642 |
| Change rest to post peak exercise | -0.30 (-0.46 to -0.15) | -0.17 (-0.32 to -0.02) | 0.13 (-0.08 to 0.35) | -0.28 (-0.43 to -0.14) | -0.22 (-0.37 to -0.07) | 0.06 (-0.14 to 0.27) | -0.07 (-0.37 to 0.23) | 0.741 |

Values presented as mean ± SE or mean difference (95% CI). AZA = Acetazolamide, bpm = beats per minute, CI = 95% confidence interval, HR = heart rate, SpO<sub>2</sub> = oxygen saturation measured by pulse oximetry, SD = Standard deviation, PaCO<sub>2</sub> = partial pressure of carbon dioxide in arterial blood, PaO<sub>2</sub> = partial pressure of oxygen in arterial blood.

At 760 m and J + 60, we observed that 3 (6%) patients assigned to placebo and 3 (6%) patients assigned to acetazolamide had STE of ≥0.3 mm in aVR during post-peak exercise (Figure 2, Panel B). At J + 80, none and 2 (4%) of the patients assigned to placebo and acetazolamide, respectively, exceeded the STE threshold of ≥0.3 mm at 760 m (Figure, Panel A) during post-peak exercise. During post-peak exercise at 3100 m under placebo and J + 60, 2 (4%) new STE ≥0.3 mm appeared and 1 (2%) STE disappeared compared to 760 m (P = 0.564, McNemar Test). With acetazolamide, 1 (2%) new STE ≥0.3 mm appeared and 2 (4%) disappeared compared to 760 m (P = 0.564, McNemar Test). No treatment effect was detected (P = 0.242, Fisher Exact Test). Correspondingly, at J + 80 under placebo, 2 (4%) new STE (P = 0.157, McNemar Test); under acetazolamide, 1 (2%) new STE and 1 (2%) absent STE was observed compared to 760 m (P = 1.000 McNemar Test). No effect of acetazolamide treatment was detected (P = 0.242, Fisher Exact Test). Results were confirmed when comparing STE at either J + 60 or J + 80 at 3100 m versus 760 m.

**Figure 2:**
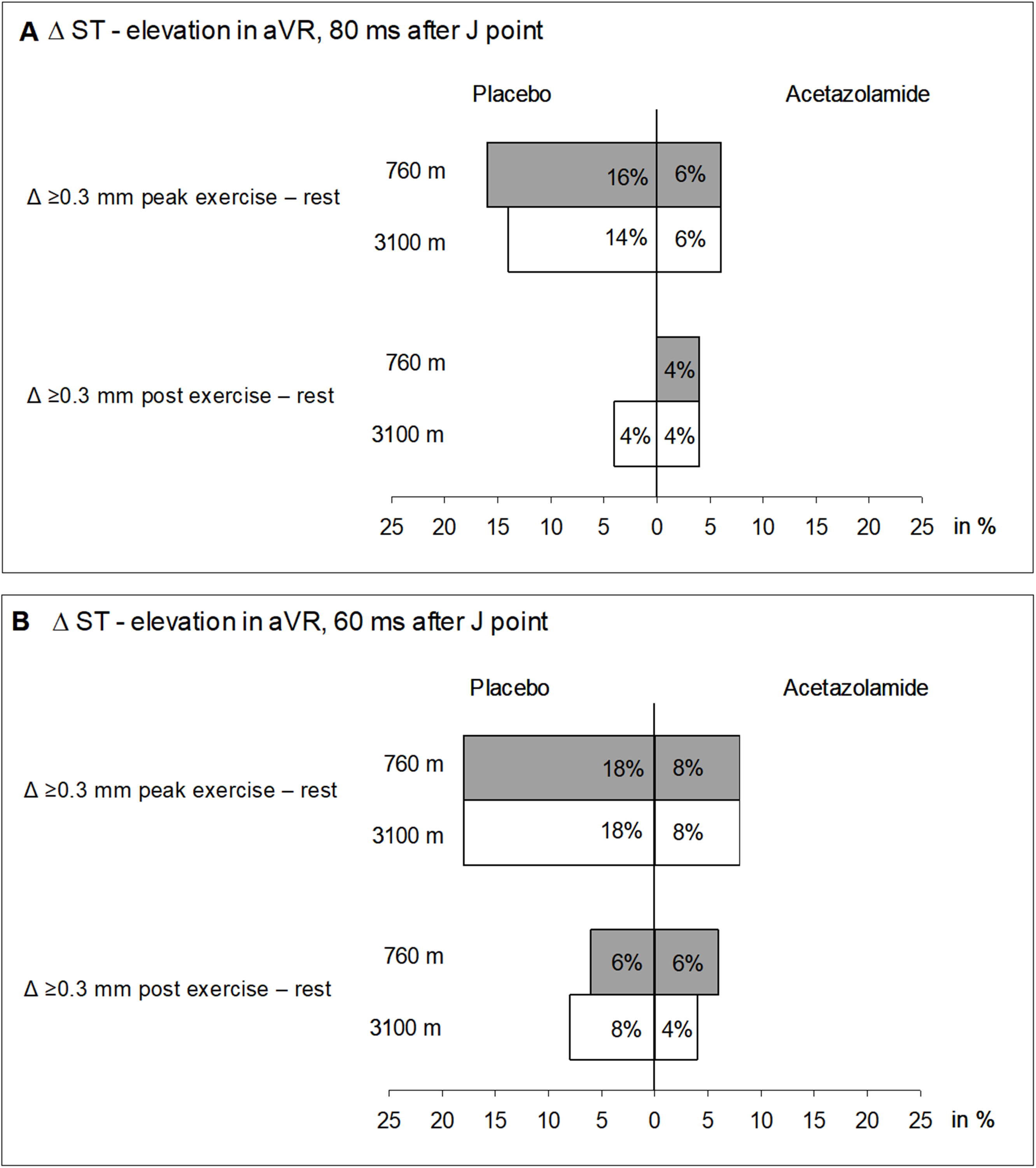
Incidence of ST-segment elevations ≥0.3 ms from values at rest measured in aVR at peak exercise and post exercise. **Panel A**, ST-segment measured from the J-point + 80 ms; **Panel B**, ST-segment measured from the J-point + 60 ms.

Additionally, no changes in the occurrence or absence of STE during rest or peak exercise between altitudes and interventions were observed.

### Cardiopulmonary exercise tests

The maximal work rate (Wmax) was significantly reduced at 3100 m compared to 760 m in both groups by a mean difference (95%CI) of 11 W (7 to 14) and 14 W (11 to 17) under placebo and acetazolamide, respectively (Table 2). Acetazolamide showed no significant treatment effect regarding Wmax in comparison to placebo. Arterial blood gas analysis at peak exercise revealed significant changes with ascent from 760 to 3100 m, i.e., SpO_2_, mean difference of −9.7% (−10.5 to −9.0), PaO_2_ of −2.5 kPa (−2.7 to −2.3) and PaCO_2_ of −0.4 kPa (−0.5 to −0.3). Acetazolamide significantly mitigated altitude-related decrements in SpO_2_ and PaO_2_ at peak exercise by 2.0% (1.0 to 3.1) and 0.8 kPa (0.5 to 1.1), respectively, whereas PaCO_2_ was further reduced by −0.3 kPa (− 0.5 to −0.2) (Table 2).

68 of 90 (76 %) of patients randomized to placebo experienced various ARAHE during the stay at 3100 m while the incidence of ARAHE in the acetazolamide group was 42 of 86 (49 %), p<0.001, chi-squared test.

## Discussion

We analyzed ST-segment changes in aVR as signs of cardiac ischemia at rest, at peak and post peak exercise in patients with COPD during a sojourn at 3100 m and treated with either placebo or acetazolamide. We found that about 6% lowlanders with moderate to severe COPD did have clinically relevant STE at 760 m, but no further worsening during the first hours at 3100 m was observed. This percentage was similar in COPD-patients ascending to 3100 m under acetazolamide prophylaxis. Although, patients with COPD experienced various ARAHE during their stay at 3100 m that were partially prevented by acetazolamide,(11) no clinically relevant ECG abnormalities were associated. During their stay at 3100 m patients with COPD experienced various ARAHE that were partially prevented by acetazolamide, although no clinically relevant ECG abnormalities were associated.

Data on ECG-changes upon exposure to altitude in the general population and patients with COPD are scant. Only two related studies by our research group were identified in a detailed search of the literature.(9, 10) The study by Bisang et al. reported prolonged QT-intervals corrected for heart rate in COPD patients staying at 2048 m, which were not prevented by nocturnal oxygen therapy.(9) ST changes were not reported. In a previous study of our group we applied 12-lead ECG in patients with moderate to severe COPD that traveled to 3100 m and exercised after the same time spent at the target altitude.(10) This study confirms our findings that altitude-exposure did not induce ST-segment changes in V5. It is reassuring that neither the current nor the cited study by Carta et al., found new STE at rest or peak exercise at 3100 m.

Apart from confirming the findings from Carta et al., the current study reports for the first time ECG data from post exercise. Notably, STE in aVR during post exercise had the highest prognostic values for predicting myocardial ischemia in a previous study by Wagener et al.(8) In the study, 1596 patients, of whom an exercise inducible cardiac ischemia was suspected were examined. ST-segment amplitudes in leads aVR and V5 were measured automatically. As a reference as to whether the patient actually had an ischemia or not, myocardial perfusion-single photon emission computed tomography (MP-SPECT) was used. Maximal accuracy (area under the curve = 0.62) was achieved in lead aVR with a cut-off value of 0.030 mV (equal to 0.3 mm) for STE at 2 min into recovery and 80 ms after the J point. The results further showed that in patients with cardiac ischemia, the STE was significantly higher than in non-ischemic patients in lead aVR as well as V5. The diagnostic accuracy for ischemia of lead aVR alone was equally high as lead V5.

Accordingly, due to simplicity and worldwide feasibility of applying 4-lead ECG, we decided in our study to focus exclusively on lead aVR and chose the same cut-off value of 0.3 mm. Despite of the measurements taken at J + 60 or J + 80 ms, our study was not able to show a significant change in ST-segments due to altitude or acetazolamide. Although, the statistically significant increase in the STE (at J + 60 and J + 80 ms) from rest to post-exercise at 3100 m compared to 760 m under placebo suggesting provocation of some degree of cardiac ischemia, the clinical relevance of this finding remains to be elucidated.

Our study is the first to estimate the prevalence of STE in COPD patients at rest and during exercise at low and high altitude. McKinney and Pitcher(16) tried to determine the prevalence of STE in aVR in a population of over 2000 patients referred for exercise ECG recordings for evaluation of the diagnosis of a coronary artery disease. They automatically measured the STE at 80 ms after the J point and used a cut-off value of STE ≥ 1 mm. All STEs above 0.75 mm were additionally checked by a cardiologist, which may have increased the accuracy of this study. Their results showed that 3.4% of the patients had a STE in aVR. In the current study, we found low proportions of post-exercise STE of 0 to 6% at J + 60 and J + 80 ms, respectively (Figure 2).

A decrease in SpO_2_, PaO_2_ and PaCO_2_ at peak exercise at high altitude has previously been observed in COPD patients.(17, 18) Our findings also confirm the results of Jonk et al.(19) She observed blood gas changes in healthy individuals during exercise in normoxic and hypoxic conditions under placebo and acetazolamide treatment. The hyperventilation induced by acetazolamide is consistent with the improvements in SpO_2_ and PaO_2_, however, acetazolamide did not improve exercise performance. Indeed, the effect of acetazolamide on maximal exercise performance at high altitude remains under debate and needs further investigations.(20)

The current study investigated STE changes in non-severely hypoxemic, stable COPD patients without serious or unstable cardiopulmonary comorbidities. Therefore, this population is representative of a large community of COPD patients but not of patients with pronounced hypoxemia or hypercapnia at lowlands. Furthermore, results should not be extrapolated to patients with more severe COPD. Exercise testing and the administration of acetazolamide was well tolerated. The focus of the current study was to investigate STE changes in aVR, which can be assessed by a simple 4-lead ECG recording. However, this limits the interpretation, since ST changes in inferior leads could not be detected.

## Conclusion

This study in non-severely hypoxemic, stable lowlanders with moderate to severe COPD without any serious cardiopulmonary comorbidities confirms previous findings that a short altitude sojourn at 3100 m is associated with statistically significant but clinically most likely not relevant ECG changes in aVR. This study adds new valuable information regarding STE changes in aVR during the post-exercise period, a condition not yet reported in COPD patients at high altitude, which was not modified by acetazo-lamide prophylaxis.

## Data Availability

All data produced in the present study are available upon reasonable request to the authors.

## Acknowledgments

Siemens Health Engineers provided some equipment for the study.

## Funding

The study was supported by the Swiss National Science Foundation (172980) and Lunge Zurich.

## Disclosure

The authors declare no conflict of interest.

## Data availability statement

Anonymized data underlying this study can be requested by qualified researchers providing an approved proposal.

## Notes

### Competing Interest Statement

The authors have declared no competing interest.

### Clinical Trial

NCT03156231

### Author Declarations

Ethics Committee of the National Center of Cardiology and Internal Medicine, Bishkek, Kyrgyzstan, gave ethical approval for this work (08-2016).

